# The Association between Proteomic Aging Clocks and the Risk of Cancer in Midlife Individuals

**DOI:** 10.1101/2025.01.05.25320018

**Authors:** Shuo Wang, Zexi Rao, Aixin Li, Anne H. Blaes, Michael J. Blaha, Josef Coresh, Ruth Dubin, Rajat Deo, Corinne E. Joshu, Catherine Handy Marshall, James S. Pankow, Jerome I. Rotter, Bharat Thyagarajan, Seamus P. Whelton, Peter Ganz, Weihua Guan, Elizabeth A. Platz, Anna Prizment

## Abstract

**Background:** To measure the aging process before a cancer diagnosis, we developed the first cancer-specific proteomic aging clock (CaPAC) and examined its association with cancer risk in the Atherosclerosis Risk in Communities (ARIC) and Multi-Ethnic Study of Atherosclerosis (MESA) studies.

**Methods:** Using the SomaScan assay, ARIC measured 4,712 proteins in plasma samples collected in 1990-92 from 3,347 participants who developed cancer over follow-up until 2015 and 7,487 who remained cancer-free, all aged 46-70. We constructed CaPAC0 using elastic net regression among two-thirds randomly selected cancer-free participants (N=4,991, training set) and calculated age acceleration for CaPAC0 (CaPAA0) as residuals of CaPAC0 on chronological age in all remaining ARIC participants. We used multivariable-adjusted Cox proportional hazards regression to calculate hazard ratios (HRs) for the risk of overall, obesity-related, smoking-related, and the most common cancers (prostate, lung, breast, colorectal) with CaPAA0 using a case-cohort design. We replicated the analysis in 3,893 MESA participants aged 46-70 at Exam 1 (456 incident cancer).

**Results:** CaPAC0 was correlated with chronological age in ARIC and MESA (r=0.82 and 0.86, respectively). In both ARIC and MESA, CaPAA0 was significantly (p<0.05) associated with the risk of overall [HRs per 5-years=1.08 and 1.23, respectively], smoking-related [HRs=1.30 and 1.54, respectively], and lung cancers [HRs=1.54 and 1.94, respectively]. CaPAA0 was also significantly associated with colorectal cancer risk in ARIC [HR=1.31], but not in MESA. CaPAA0 was not associated with obesity-related, breast, or prostate cancers.

**Conclusion:** CaPAA0 was associated with several types of cancer with the strongest association observed for lung cancer risk.

## Introduction

Cancer patients experience accelerated aging, but it is unclear whether accelerated aging starts before their cancer diagnosis. To measure the aging process before cancer diagnosis, researchers have proposed biological age estimators called aging clocks, such as epigenetic clocks (based on DNA methylation sites) and more novel proteomic aging clocks (PACs, based on circulating proteins).^1^ Epigenetic clocks have been associated with an increased risk of overall,^2–4^ lung,^5^ breast,^6,7^ and colorectal cancers,^3^ but estimates differed by study and clock. Moreover, the biological functions of DNA methylation sites are unclear.^8^ Thus, we examined PACs,^9–12^ because proteins, as intermediates between genetics and disease, may provide information on age-related pathologies.^13,14^ Additionally, 96% of FDA-approved drug targets are proteins;^15^ therefore, the proteins comprising PACs may be targets for anti-aging drugs. However, no studies have developed a cancer-specific PAC (CaPAC) and examined its association with cancer risk.

We developed CaPACs in the Atherosclerosis Risk in Communities (ARIC) study, a population-based cohort of White and Black females and males, who were followed for cancer from 1987-2015. Using the SomaScan® 5K assay, ARIC measured 4,712 proteins in plasma samples from 1990-92 (aged 46-70, midlife) and 2011-13 (aged 66-90, late life). We first developed CaPAC0 among midlife participants who remained cancer-free until 2015 and examined its association with the risk of overall, obesity-related, smoking-related, and the most common cancers (breast, prostate, lung, colorectal) in ARIC and replicated the analysis in the Multi-Ethnic Study of Atherosclerosis (MESA) study. Moreover, we developed two additional CaPACs: 1) CaPAC1, developed similarly as CaPAC0 but limited to ∼20 proteins; and 2) CaPAC2, which included 420 proteins with the largest within-individual changes over 20 years (midlife to late life). We also computed the published Lehallier’s PAC developed in predominantly White healthy individuals (Lehallier [2020]).^9^ We compared the associations of CaPAC1, CaPAC2, and Lehallier’s PAC with cancer to those for CaPAC0. Finally, we computed the Klemera-Doubal biological age (KDM-BA), constructed using clinical and physiological markers (Levine [2013]),^16^ and examined whether CaPAC0 is associated with cancer risk independent of KDM-BA.

## Methods

A detailed methods section can be found in the **Supplemental Methods**.

### The ARIC prospective cohort study

ARIC enrolled 15,744 White and Black participants in 1987-89 from four centers (Maryland, Minnesota, Mississippi, North Carolina).^17,18^ Thus far, ten visits have been completed.^17^ Participants underwent telephone follow-up annually in 1987-2012 and semi-annually after 2012, with response rates of 83%-99% for follow-up calls among alive participants.^18^

Incident cancer was ascertained through 2015 via state cancer registries in four centers, supplemented by the abstraction of medical records and hospital discharge summaries.^18^ Obesity-related cancers included oropharynx, esophagus, postmenopausal breast, liver, gallbladder, pancreatic, stomach, kidney, colorectal, endometrial, ovarian, and lethal prostate cancers.^19^ Smoking-related cancers included head and neck, esophagus, lung, laryngeal, bladder, liver, pancreatic, stomach, kidney, colorectal, and cervical cancers.^20^

ARIC measured 4,955 aptamers (corresponding to 4,712 proteins) using SomaScan® 5K in plasma samples collected at Visit 2 (1990-92, midlife participants) and Visit 5 (2011-13, late-life). Bland-Altman coefficient of variation for split samples was 6% and 7% at Visits 2 and 5, respectively.

### The MESA prospective cohort study

MESA enrolled 6,814 White, Black, Hispanic, and Chinese participants in 2000-02 (Exam 1) from six centers. Every 9-12 months, participants self-reported events including hospitalizations, prompting requests for hospitalization records. Hospitalization records contained International Classification of Diseases, 9th Revision (ICD-9) diagnosis including cancer codes (140-209), which were used to ascertain incident cancer until 2018.^21^ MESA measured ∼7,000 proteins in blood samples collected at Exam 1 using SomaScan® 7K, which included all proteins measured in ARIC.

Both studies were approved by institutional review boards, and participants provided written informed consent. In both studies, information about demographics, lifestyle factors, medication use, and medical history has been collected at each visit; protein concentrations, expressed in relative fluorescent units, were log2-transformed to correct for skewness.

### Statistical analysis

CaPACs were trained against chronological age using elastic net regression (alpha=0.5, lambda selected by 10-fold cross-validation)^10,22,23^ in R (version 4.1.2, package “glmnet”). All the other analyses were performed using SAS version 9.4 (SAS Institute Inc, Cary, NC). P-value<0.05 for two-sided tests was considered statistical significance.

Among midlife participants (aged 46-70) attending ARIC Visit 2 (1990-92), 3,347 developed cancer through 2015, while 7,487 remained free of a cancer diagnosis (cancer-free participants) (**Figure 1**). We randomly selected two-thirds of cancer-free participants (training set, N=4,991) and constructed CaPAC0 as a weighted sum of 1,282 aptamers selected by elastic net regression. We described the functions of the top 20 proteins with the largest absolute weights in CaPAC0 using GeneCards (https://www.genecards.org/).

**Figure 1.**
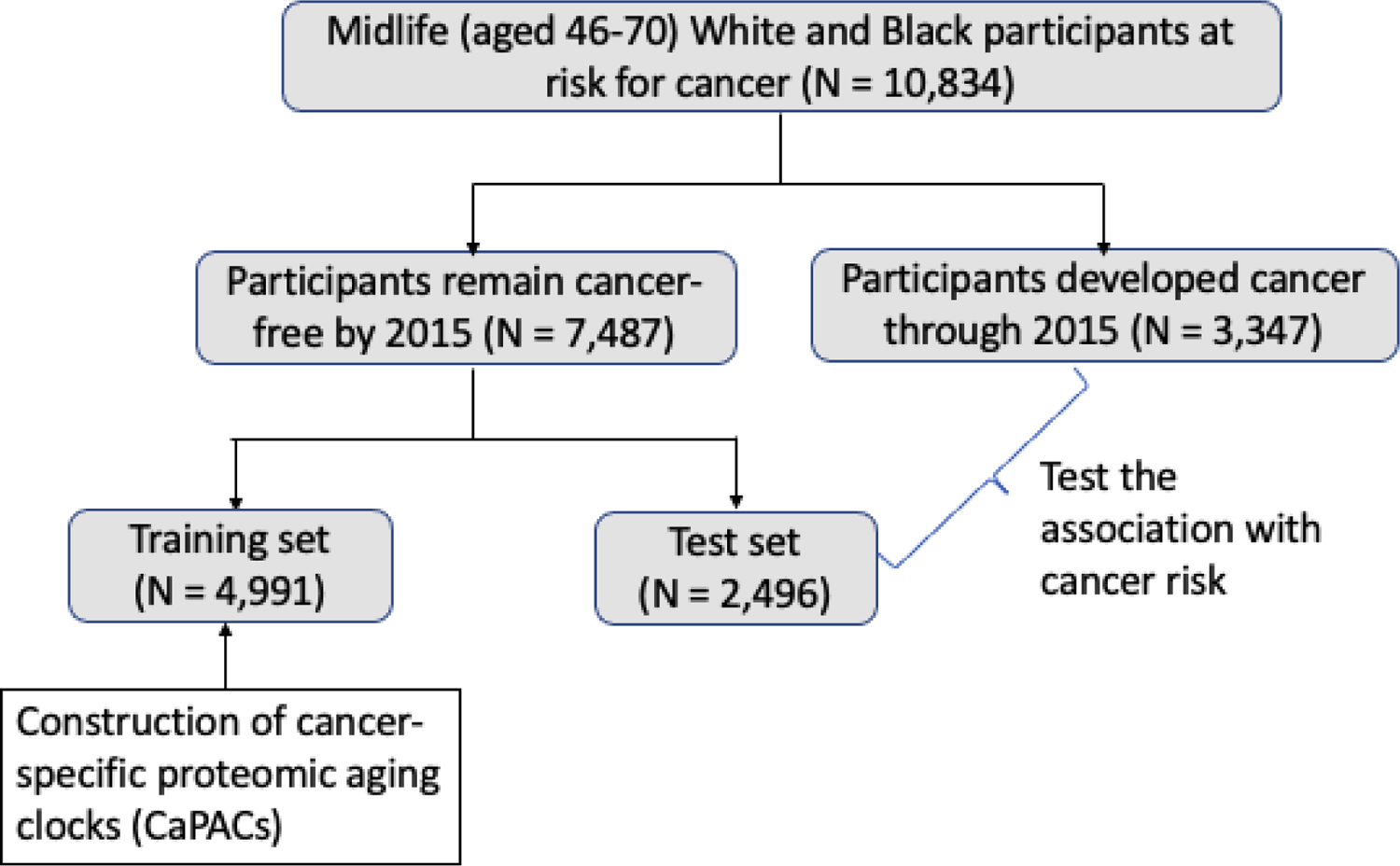
Study population for the development of CaPACs, and for testing the association of the CaPACs with cancer risk in ARIC

We calculated Pearson correlation between CaPAC0 and chronological age and median absolute error (MAE). To capture CaPAC0’s association with cancer independent of chronological age, we estimated age acceleration for CaPAC0 (CaPAA0) as the residuals after regressing CaPAC0 on chronological age. We compared the distributions of characteristics among cancer-free participants (test set) and those who developed cancer. We used Cox proportional hazards regression to estimate hazard ratios (HRs) and 95% confidence intervals (CIs) for the risk of overall, obesity-related, smoking-related, and the most common cancers, i.e., breast (postmenopausal in this study), prostate, lung, and colorectal, per 5-year increase in CaPAA0. We modeled CaPAA0 as a continuous variable because no nonlinear association was observed when applying cubic splines. Total person-years were determined from Visit 2 until cancer diagnosis, death, loss to follow-up, or censoring on 2015/12/31, whichever occurred first. Proportional hazards assumption was examined by graphical methods and was not violated in any models.

To account for the exclusion of training set used to construct CaPAC0, we applied a case-cohort study design employing Barlow’s method (**Supplemental Methods).**^24^ We constructed two models: a model adjusted for demographics (chronological age, sex, race-center) and a model further adjusted for cancer-related risk factors, such as education, BMI, smoking status, pack-years of smoking, alcohol intake, aspirin use, hormone replacement therapy use (among females) and diabetes, and for eGFR because it was associated with protein levels (fully-adjusted model, see **Supplemental Methods**). We reported results only for a fully-adjusted model because HRs for all cancers were similar in the models.

Participants with missing covariates of interest were excluded from the analysis of CaPAA0 and cancer. To exclude the effect of undiagnosed cancer on plasma proteins, in a sensitivity analysis, we excluded participants who developed cancer within two and four years of blood collection and replicated the analysis for cancers significantly associated with CaPAA0 (overall, smoking-related, lung, and colorectal cancers) in ARIC.

At MESA Exam 1 (2000-02), we calculated CaPAC0 using proteins and weights from ARIC in 3,893 cancer-free participants in the subset aged 46-70 years (to have the same age range as in ARIC). We examined the association between CaPAA0 and incident cancer (N=456) until 2018. Additionally, we applied CaPAC0 to all cancer-free participants (45-84 years) at Exam 1 (N=5,330) and examined its association with incident cancer (N=678).

In ARIC and MESA, we also examined associations of CaPAA0 with overall and smoking-related cancers stratified by sex, race, and smoking status, because males^25^ and smokers^4^ might age faster and Black individuals had a higher incidence of several cancer types.^26^ In ARIC, we conducted two additional analyses: 1) we excluded one common cancer at a time to explore whether the associations with smoking-related cancers were driven by those cancers and 2) we stratified the association between CaPAA0 and lung cancer by smoking status, because smoking is a strong risk factor for lung cancer. These analyses were not conducted in MESA due to the limited sample size.

We developed two additional CaPACs. First, we developed CaPAC1, consisting of 21 aptamers, because measuring a CaPAC with fewer proteins (∼20 aptamers) would be cost-effective and applicable in clinics in the future. We examined if CaPAC1 shows similar associations with cancer risk as those for the CaPAC0 consisting of 1,282 aptamers. Second, to better capture proteins that change with age, we selected the top 40% of proteins with the largest change from midlife to late life (over 20 years) and constructed CaPAC2, which included 420 aptamers selected by elastic net regression. For comparison, we computed the published Lehallier’s PAC, created in healthy White individuals (**Supplemental Methods**), and tested its association with cancer risk. Finally, in ARIC, we computed KDM-BA^16^ (**Supplemental Methods**), constructed using clinical and physiological markers rather than proteins measured using SomaScan. We included age acceleration for KDM-BA and CaPAA0 in the same model to examine whether CaPAC0 provides additional information about cancer risk beyond KDM-BA. For this analysis, we examined only lung cancer because age acceleration for KDM-BA was significantly associated with only lung cancer in ARIC.

## Results

CaPAC0 was correlated with chronological age in the training (r=0.92, MAE=1.57 years) as well as in ARIC (after excluding the training set, r=0.82, MAE=2.22) and MESA participants (46-70 years) (r=0.86, MAE=4.01) (**Figure 2**). Top 20 proteins with the largest absolute weights in CaPAC0 were involved in inflammation, metalloproteinase secretion, or cell proliferation (**Supplemental Table 1**).

**Figure 2.**
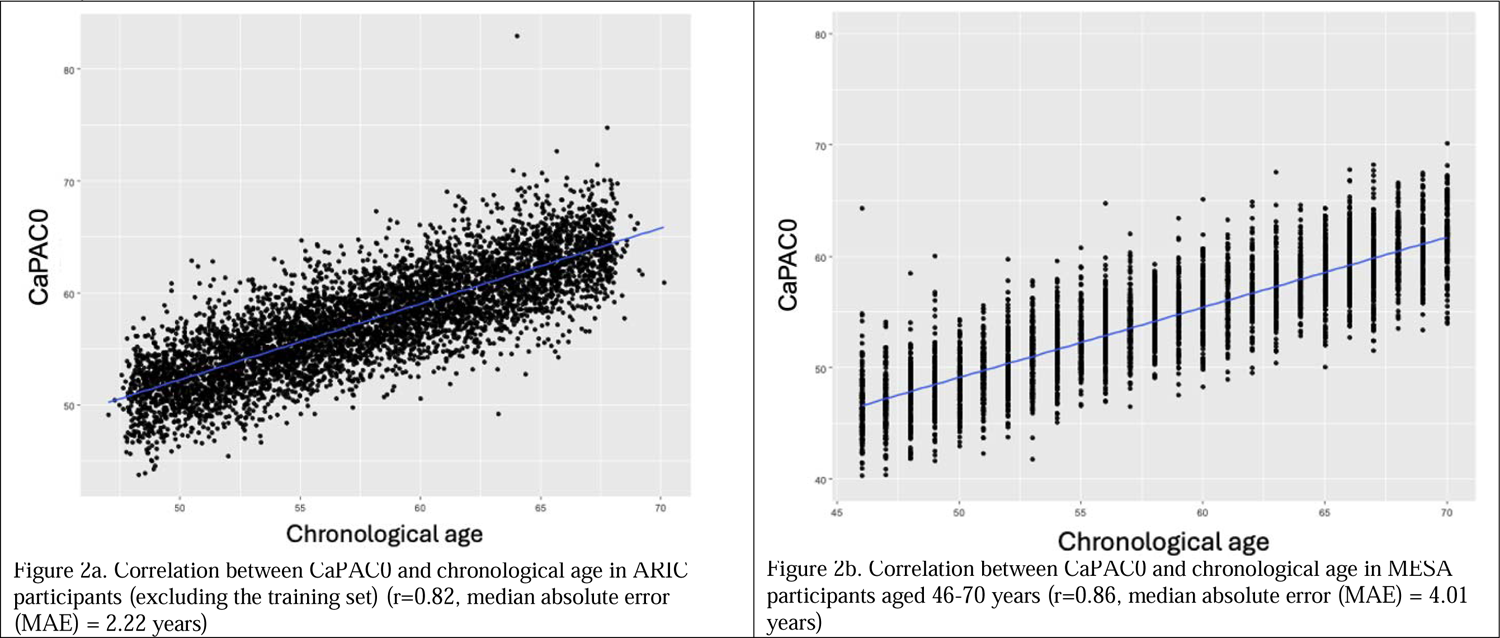
Correlation between CaPAC0 and chronological age in ARIC participants (excluding the training set), and in MESA participants aged 46-70 years

In ARIC and MESA (aged 46-70), compared to cancer-free participants, those who developed cancer tended to be current smokers and have a higher number of pack-years smoked, and were less likely to be never drinkers (**Table 1**). Additionally, participants who developed cancer in MESA (but not in ARIC) were more likely to be aspirin users and women were less likely to be ever users of hormone replacement therapy (**Table 1**).

**Table 1.** Distributions of characteristics in participants who remained cancer-free (i.e., cancer-free participants) and those who developed cancer during follow-up in ARIC and in MESA.

|  | ARIC participants (after excluding the training set, aged 46-70 years) |  |  | MESA participants aged 46-70 years |  |  |
| --- | --- | --- | --- | --- | --- | --- |
|  | Cancer-free participants<br>(test set) | Participants<br>developed cancer | P-value<sup>c</sup> | Cancer-free<br>participants | Participants<br>developed cancer | P-value |
| N of participants<sup>a</sup> | 2,172 | 2,947 |  | 3,437 | 456 |  |
| Mean age, years (SD) | 57.4 (5.7) | 58.1 (5.7) | <0.001 | 57.8 (7.3) | 60.3 (6.7) | <0.001 |
| Male, % | 39.9 | 54.1 | <0.001 | 52.4 | 44.0 | <0.001 |
| Race/Ethnicity<sup>b</sup>, % |  |  |  |  |  |  |
| White | 75.4 | 76.2 | 0.53 | 36.4 | 42.0 | <0.001 |
| Black | 24.7 | 23.8 |  | 26.4 | 32.5 |  |
| Hispanic | NA | NA |  | 24.5 | 17.7 |  |
| Chinese | NA | NA |  | 12.7 | 7.8 |  |
| Education |  |  |  |  |  |  |
| Less than high school | 22.2 | 23.3 | 0.62 | 17.2 | 12.7 | 0.05 |
| High school equivalent | 42.0 | 41.5 |  | 17.5 | 19.2 |  |
| Greater than high school | 35.8 | 35.2 |  | 65.4 | 68.1 |  |
| Mean BMI, kg/m<sup>2</sup> (SD) | 28.0 (5.5) | 28.0 (5.3) | 0.94 | 28.7 (5.6) | 29.1 (5.6) | 0.11 |
| Smoking status, % |  |  |  |  |  |  |
| Never smoker | 42.7 | 34.0 | <0.001 | 50.8 | 39.9 | <0.001 |
| Former smoker | 37.4 | 28.1 |  | 35.2 | 38.6 |  |
| Current smoker | 19.9 | 37.9 |  | 14.3 | 21.6 |  |
| Mean pack-years of smoking among<br>ever smokers (SD) | 26.3 (21.1) | 32.0 (22.9) | <0.001 | 10.3 (19.8) | 16.9 (26.2) | <0.001 |
| Alcohol intake |  |  |  |  |  |  |
| Current drinker | 55.5 | 59.7 | <0.001 | 56.0 | 57.3 | 0.007 |
| Former drinker | 20.0 | 20.4 |  | 23.7 | 28.0 |  |
| Never drinker | 24.5 | 19.9 |  | 20.3 | 14.7 |  |
| Diabetes, % | 16.5 | 14.1 | 0.02 | 25.9 | 28.2 | 0.30 |
| Aspirin use in the preceding two<br>weeks, % | 51.3 | 51.3 | 0.99 | 27.5 | 36.0 | <0.001 |
| Ever users of hormone replacement<br>therapy among females, % | 44.8 | 45.1 | 0.55 | 61.5 | 55.4 | 0.008 |
| Mean eGFR, mL/min/1.73 m<sup>2</sup> (SD) | 98.7 (17.2) | 97.8 (15.9) | 0.06 | 76.6 (16.3) | 76.7(16.7) | 0.87 |

<sup>a</sup> Participants with missing covariates of interest were excluded from the analysis.
<sup>b</sup> ARIC includes only White and Black participants, while MESA includes White, Black, Hispanic, and Chinese participants.
<sup>c</sup> P-values were calculated from t-test for continuous variables and chi-square test for categorical variables.

In ARIC and MESA (aged 46-70, median follow-up=17.19 and 17.16 years, respectively), CaPAA0 was statistically significantly associated with the risk of overall [HRs per 5-years=1.08 and 1.23, respectively] and smoking-related cancers [HRs=1.30 and 1.54, respectively], but not obesity-related cancers (**Table 2**). CaPAA0 was also associated with lung cancer in both ARIC and MESA (aged 46-70) [HRs=1.54 and 1.94, respectively] (**Table 2**). Additionally, CaPAA0 was associated with colorectal cancer in ARIC [1.31 (1.05-1.63)], but not in MESA (aged 46-70) (**Table 2**). In ARIC, after excluding those with lung or colorectal cancers (one at a time), the association of CaPAA0 with smoking-related cancers remained significant (**Supplemental Table 2**). Further, after excluding participants who developed cancer within two and four years after the protein measurement, the associations of CaPAA0 with overall, smoking-related, lung, and colorectal cancers were similar to those in the main analysis (**Supplemental Table 3**). These analyses were not conducted in MESA due to the limited sample size. CaPAA0 was not associated with postmenopausal breast or prostate cancers in participants aged 46-70 from either study (**Table 2**). However, CaPAA0 was significantly associated with prostate cancer in all MESA participants aged 45-84 years (**Supplemental Table 4**). CaPAA0’s associations with other cancer types were similar in MESA participants aged 45-84 years and those aged 46-70 years (same range as in ARIC) (**Supplemental Table 4**).

**Table 2.**
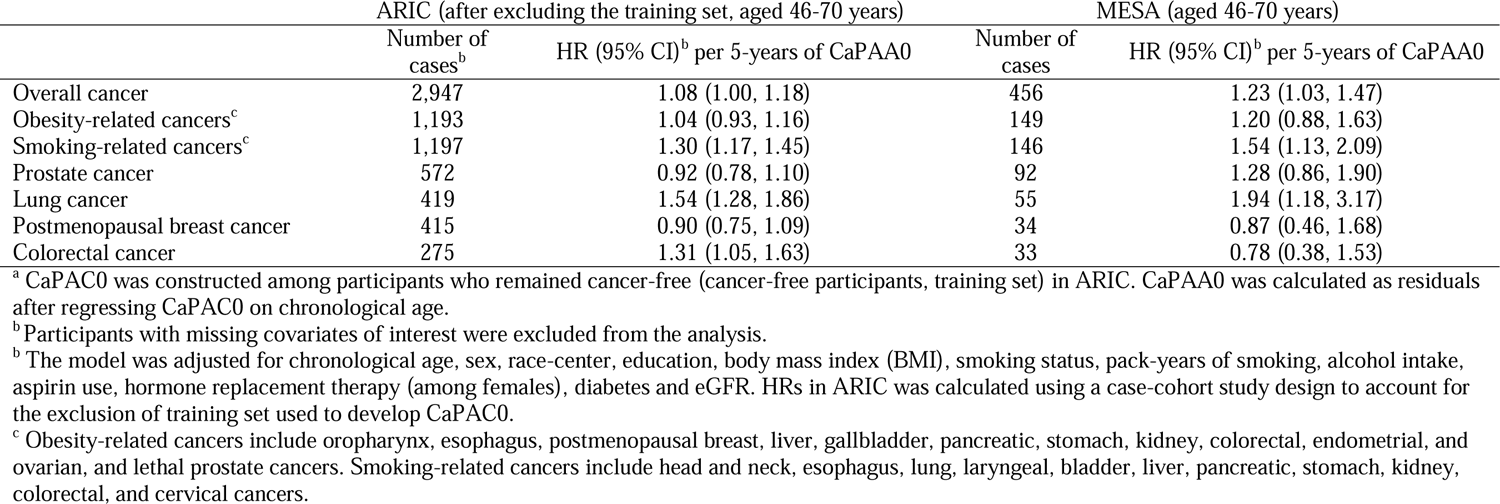
Association between age acceleration for CaPAC0 (CaPAA0)^a^ and cancer risk in ARIC (1990-2015) and in MESA (2000-2018)

|  | ARIC (after excluding the training set, aged 46-70 years) |  | MESA (aged 46-70 years) |  |
| --- | --- | --- | --- | --- |
|  | Number of cases<sup>b</sup> | HR (95% CI)<sup>b</sup> per 5-years of CaPAA0 | Number of cases | HR (95% CI)<sup>b</sup> per 5-years of CaPAA0 |
| Overall cancer | 2,947 | 1.08 (1.00, 1.18) | 456 | 1.23 (1.03, 1.47) |
| Obesity-related cancers<sup>c</sup> | 1,193 | 1.04 (0.93, 1.16) | 149 | 1.20 (0.88, 1.63) |
| Smoking-related cancers<sup>c</sup> | 1,197 | 1.30 (1.17, 1.45) | 146 | 1.54 (1.13, 2.09) |
| Prostate cancer | 572 | 0.92 (0.78, 1.10) | 92 | 1.28 (0.86, 1.90) |
| Lung cancer | 419 | 1.54 (1.28, 1.86) | 55 | 1.94 (1.18, 3.17) |
| Postmenopausal breast cancer | 415 | 0.90 (0.75, 1.09) | 34 | 0.87 (0.46, 1.68) |
| Colorectal cancer | 275 | 1.31 (1.05, 1.63) | 33 | 0.78 (0.38, 1.53) |

<sup>a</sup> CaPAC0 was constructed among participants who remained cancer-free (cancer-free participants, training set) in ARIC. CaPAA0 was calculated as residuals after regressing CaPAC0 on chronological age.
<sup>b</sup> Participants with missing covariates of interest were excluded from the analysis.
<sup>b</sup> The model was adjusted for chronological age, sex, race-center, education, body mass index (BMI), smoking status, pack-years of smoking, alcohol intake, aspirin use, hormone replacement therapy (among females), diabetes and eGFR. HRs in ARIC was calculated using a case-cohort study design to account for the exclusion of training set used to develop CaPAC0.
<sup>c</sup> Obesity-related cancers include oropharynx, esophagus, postmenopausal breast, liver, gallbladder, pancreatic, stomach, kidney, colorectal, endometrial, and ovarian, and lethal prostate cancers. Smoking-related cancers include head and neck, esophagus, lung, laryngeal, bladder, liver, pancreatic, stomach, kidney, colorectal, and cervical cancers.

In stratified analyses, CaPAA0’s association with overall, smoking-related, and lung cancers did not differ by smoking status; the HRs among never smokers remained increased but some associations lost statistical significance (**Table 3**). The association between CaPAA0 and overall cancer was higher in males than females in MESA (p-interaction=0.03) but not in ARIC. CaPAA0 had similar associations with overall cancer, respectively, among White and Black participants in ARIC and MESA (**Table 3**); however, in MESA, HRs for overall cancer were lower among White and Black than among Hispanic and Chinese participants (**Table 3**). A similar pattern of HRs across race/ethnicity groups was observed for smoking-related cancers (**Table 3**).

**Table 3.**
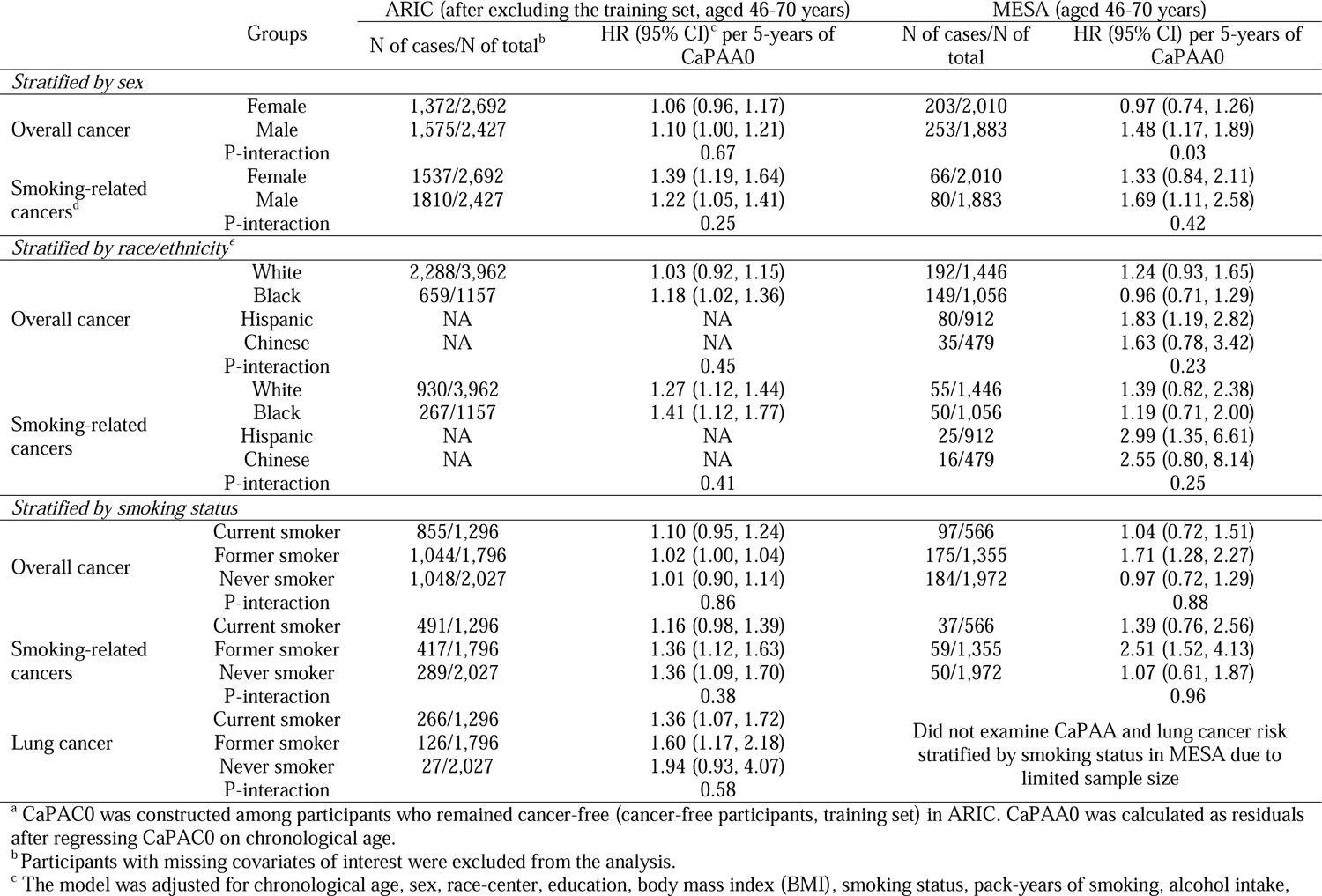
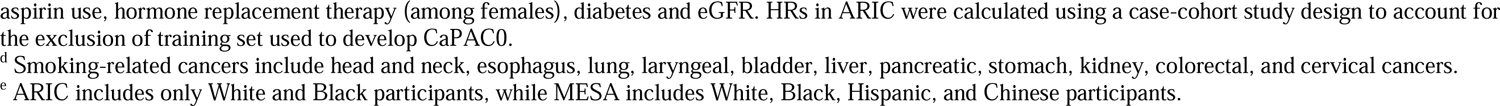
Association between age acceleration for CaPAC0 (CaPAA0)^a^ and cancer risk stratified by sex, race/ethnicity, and smoking status among participants in ARIC (1990-2015) and in MESA (2000-2018)

| Groups |  | ARIC (after excluding the training set, aged 46-70 years) |  | MESA (aged 46-70 years) |  |
| --- | --- | --- | --- | --- | --- |
|  |  | N of cases/N of total<sup>b</sup> | HR (95% CI)<sup>c</sup> per 5-years of CaPAA0 | N of cases/N of total | HR (95% CI) per 5-years of CaPAA0 |
| *Stratified by sex* |  |  |  |  |  |
| Overall cancer | Female | 1,372/2,692 | 1.06 (0.96, 1.17) | 203/2,010 | 0.97 (0.74, 1.26) |
|  | Male | 1,575/2,427 | 1.10 (1.00, 1.21) | 253/1,883 | 1.48 (1.17, 1.89) |
|  | P-interaction |  | 0.67 |  | 0.03 |
| Smoking-related cancers<sup>d</sup> | Female | 1537/2,692 | 1.39 (1.19, 1.64) | 66/2,010 | 1.33 (0.84, 2.11) |
|  | Male | 1810/2,427 | 1.22 (1.05, 1.41) | 80/1,883 | 1.69 (1.11, 2.58) |
|  | P-interaction |  | 0.25 |  | 0.42 |
| *Stratified by race/ethnicity<sup>e</sup>* |  |  |  |  |  |
| Overall cancer | White | 2,288/3,962 | 1.03 (0.92, 1.15) | 192/1,446 | 1.24 (0.93, 1.65) |
|  | Black | 659/1157 | 1.18 (1.02, 1.36) | 149/1,056 | 0.96 (0.71, 1.29) |
|  | Hispanic | NA | NA | 80/912 | 1.83 (1.19, 2.82) |
|  | Chinese | NA | NA | 35/479 | 1.63 (0.78, 3.42) |
|  | P-interaction |  | 0.45 |  | 0.23 |
| Smoking-related cancers | White | 930/3,962 | 1.27 (1.12, 1.44) | 55/1,446 | 1.39 (0.82, 2.38) |
|  | Black | 267/1157 | 1.41 (1.12, 1.77) | 50/1,056 | 1.19 (0.71, 2.00) |
|  | Hispanic | NA | NA | 25/912 | 2.99 (1.35, 6.61) |
|  | Chinese | NA | NA | 16/479 | 2.55 (0.80, 8.14) |
|  | P-interaction |  | 0.41 |  | 0.25 |
| *Stratified by smoking status* |  |  |  |  |  |
| Overall cancer | Current smoker | 855/1,296 | 1.10 (0.95, 1.24) | 97/566 | 1.04 (0.72, 1.51) |
|  | Former smoker | 1,044/1,796 | 1.02 (1.00, 1.04) | 175/1,355 | 1.71 (1.28, 2.27) |
|  | Never smoker | 1,048/2,027 | 1.01 (0.90, 1.14) | 184/1,972 | 0.97 (0.72, 1.29) |
|  | P-interaction |  | 0.86 |  | 0.88 |
| Smoking-related cancers | Current smoker | 491/1,296 | 1.16 (0.98, 1.39) | 37/566 | 1.39 (0.76, 2.56) |
|  | Former smoker | 417/1,796 | 1.36 (1.12, 1.63) | 59/1,355 | 2.51 (1.52, 4.13) |
|  | Never smoker | 289/2,027 | 1.36 (1.09, 1.70) | 50/1,972 | 1.07 (0.61, 1.87) |
|  | P-interaction |  | 0.38 |  | 0.96 |
| Lung cancer | Current smoker | 266/1,296 | 1.36 (1.07, 1.72) | Did not examine CaPAA and lung cancer risk stratified by smoking status in MESA due to limited sample size |  |
|  | Former smoker | 126/1,796 | 1.60 (1.17, 2.18) |  |  |
|  | Never smoker | 27/2,027 | 1.94 (0.93, 4.07) |  |  |
|  | P-interaction |  | 0.58 |  |  |

<sup>a</sup> CaPAC0 was constructed among participants who remained cancer-free (cancer-free participants, training set) in ARIC. CaPAA0 was calculated as residuals after regressing CaPAC0 on chronological age.
<sup>b</sup> Participants with missing covariates of interest were excluded from the analysis.
<sup>c</sup> The model was adjusted for chronological age, sex, race-center, education, body mass index (BMI), smoking status, pack-years of smoking, alcohol intake,
aspirin use, hormone replacement therapy (among females), diabetes and eGFR. HRs in ARIC were calculated using a case-cohort study design to account for the exclusion of training set used to develop CaPAC0.
<sup>d</sup> Smoking-related cancers include head and neck, esophagus, lung, laryngeal, bladder, liver, pancreatic, stomach, kidney, colorectal, and cervical cancers.
<sup>e</sup> ARIC includes only White and Black participants, while MESA includes White, Black, Hispanic, and Chinese participants.

### Developing and testing of two additional CaPACs

In parallel to CaPAC0, age acceleration for CaPAC1 was associated with overall, smoking-related, and lung cancers in both ARIC and MESA and with colorectal cancer in ARIC. Additionally, it was associated with obesity-related cancers in MESA (**Supplemental Table 5**).

In both ARIC and MESA, age acceleration for CaPAC2 (including 420 aptamers selected from top 40% of proteins with the largest change from midlife to late life) and CaPAA0 was similarly associated with smoking-related cancers. Additionally, age acceleration for CaPAC2 was positively associated with lung cancer in ARIC and with overall and obesity-related cancers in MESA, while inversely associated with postmenopausal breast cancer in ARIC (**Supplemental Table 5**).

### Comparison with Lehallier’s PAC

HRs for overall, smoking-related, and lung cancers were similar for age acceleration for Lehallier’s PAC (Lehallier’s PAA) and CaPAA0, but not all these associations for Lehallier’s PAA were significant (**Supplemental Table 6**). Additionally, similarly to CaPAA0 in all MESA participants (aged 45-84 years), Lehallier’s PAA was associated with prostate cancer risk in MESA, but not in ARIC (**Supplemental Table 6**). Lehallier’s PAA was not associated with obesity-related, breast, or colorectal cancers in either study (**Supplemental Table 6**).

### Comparison with KDM-BA

The correlation between age acceleration for KDM-BA and CaPAA0 was 0.12 in ARIC. When including age acceleration for KDM-BA and CaPAA0 into the same model, CaPAA0 remained significantly associated with lung cancer risk (**Supplemental Table 7**).

## Discussion

Cancer is a disease of aging;^27–30^ most cancers develop slowly with age and have a long subclinical period, providing opportunities for prevention or interception.^31^ Proteomic aging clocks (PACs) that serve as an aging measure could provide a new method for identifying individuals at high risk of cancer. These individuals would benefit the most from frequent screening and anti-aging interventions through lifestyle changes and anti-aging therapies that are currently under active development.^32^

In our study, we developed CaPAC0, a cancer-specific PAC, in a bi-racial ARIC cohort (White: 75%; Black: 25%), and validated it in a multi-ethnic MESA cohort (White: 38%; Black: 28%; Hispanic: 22%; and Chinese: 12%). In both cohorts, CaPAC0 was strongly correlated with chronological age and age acceleration for CaPAC0 (CaPAA0) in midlife was associated with the risk of overall, smoking-related, and lung cancers in late life. In parallel, positive associations with these cancers were found for the previously published Lehallier’s PAC in ARIC and MESA, although not all these associations reached statistical significance. The association between CaPAA0 and overall cancer was similar in White and Black participants in both ARIC and MESA but were notably stronger among Hispanic and Chinese participants. A similar pattern of HRs across race/ethnicity groups was observed for smoking-related cancers. Higher hazard ratios (HRs) among Hispanic and Chinese participants could reflect true stronger associations or could be due to chance because of the limited participants’ number.

To our knowledge, our study was the first to develop a cancer-specific PAC and examined its association with cancer. Two other studies examined associations between PACs that are not cancer-specific and cancer risk: Kuo [2024] (trained against mortality)^12^ and Argentieri [2024] (trained against chronological age).^11^ Those studies were conducted in the UK Biobank study, a cohort of predominantly White (94%) individuals, and proteins were measured using a proteomic assay OLINK. Our findings for CaPAA0 and overall cancer in ARIC and MESA [HRs per 1 year=1.02-1.04] were similar to that by Kuo [2024] [HR per 1 year=1.02], whereas for lung cancer, our HRs per 1 year=1.09-1.14 were slightly stronger than those by Kuo [2024]^12^ and Argentieri [2024]^11^ [HRs per 1 year=1.05-1.07]. The parallel findings for CaPAC0 and Kuo’s PAC with overall and lung cancers are especially impressive given that Kuo’s PAC was trained against mortality, not chronological age.

Although association of CaPAA0 with the overall and lung cancers were comparable across MESA and ARIC and across different PACs, and they mirrored those in the UK Biobank, the associations with other cancers were inconsistent. CaPAA0 was associated with colorectal cancer risk in ARIC but not in MESA, and with prostate cancer in MESA (participants aged 45-84 years) but not in ARIC. These differences may be partially explained by different cancer ascertainment methods in ARIC and MESA.

While ARIC ascertained cancer through state cancer registries and supplemented by medical records, MESA used ICD-9 codes from hospitalization records. Therefore, MESA might miss cases among participants who received cancer care solely in outpatient settings.^33,34^ Moreover, several other factors could contribute to the inconsistent associations with colorectal and prostate cancers since the associations differ across PACs even within one study. For examine, in ARIC, CaPAA0 was associated with colorectal cancer but not Lehallier’s PAA. Similarly, in the UK Biobank, age acceleration for Kuo’s PAC was associated with colorectal cancer but not for Argentieri’s PAC, whereas age acceleration for Argentieri’s PAC was associated for prostate cancer, but not for KUO’s PAC.^11,12^ Several factors may contribute to these inconsistencies such as different populations used to develop PACs (cancer-free, healthy, or general population; White or non-White participants), or how PACs were trained: against chronological age or mortality.

We found parallel associations with smoking-related cancers for CaPAA0 in ARIC and MESA. Further, after excluding participants with lung or colorectal cancers (one at a time), the associations for smoking-related cancers with CaPAA0 remained significant in ARIC. Importantly, the HRs for smoking-related and lung cancers with CaPAA0 were increased among never smokers. Finally, the association between CaPAA0 and lung cancer remained after including age acceleration for KDM-BA (based on clinical biomarkers) into the same model in ARIC, which indicated that the impact of CaPAC0 on lung cancer is independent from KDM-BA. Notably, CaPAA0 was not associated with obesity-related cancers risk. Our findings suggest that our CaPAC0 may optimally capture smoking and some other environmental factors, such as second-hand smoking and air pollution associated with smoking-related cancers,^35^ but less effectively capture obesity-related factors relevant to other cancers.

Finally, we developed CaPAC1, including a small number of proteins (N=21). Compared to CaPAC0 with a larger number of proteins, the associations for CaPAC1 with several cancer types were stronger but confidence intervals were wider. CaPACs including fewer proteins are cost-effective and should be further studied as a biological age metrics applicable in clinics.

The strengths of our study include the development and validation of cancer-specific proteomic aging clocks in two prospective population studies (ARIC and MESA) that include White and non-White individuals followed for at least 18 years. Other strengths are the use of highly sensitive multiplexed SomaScan assay to measure proteins, as well as detailed information on lifestyle factors, medication use, and medical conditions in both ARIC and MESA. Additionally, cancer cases were adjudicated in ARIC. Our study also has limitations. First, the possibility of protein degradation during long-term storage in ARIC and in MESA cannot be excluded. However, in both studies, the plasma samples were frozen at −80 °C right after their collection and have never been thawed reducing the possibility of degradation.^33,36^

Second, we developed CaPAC0 among midlife individuals (46-70 years) and found that it was similarly associated with cancer risk in participants aged 46-70 and those aged 45-84 years, suggesting that CaPAC0 may be applicable to individuals over 70. However, we do not know whether CaPAC can be applied to individuals younger than 45 years.

In brief, we found parallel associations with the risk of overall, smoking-related, and lung cancers across studies and different CaPACs. The associations were mainly similar to those for non-cancer specific PACs. The strongest and most consistent association was detected for lung cancer risk for all CaPACs suggesting that CaPACs captures smoking and other environmental exposures related to smoker-related cancers.

## Data Availability Statement

The ARIC and MESA datasets are available through BioLINCC, with appropriate study approvals consistent with NIH policies. Data request forms through BioLINCC can be accessed at https://biolincc.nhlbi.nih.gov/studies/aric/ and https://biolincc.nhlbi.nih.gov/studies/mesa/.

## Acknowledgments

The authors thank the staff and participants of the ARIC study for their important contributions. Cancer data was provided by the Maryland Cancer Registry, Center for Cancer Prevention and Control, Maryland Department of Health, with funding from the State of Maryland and the Maryland Cigarette Restitution Fund. The collection and availability of cancer registry data are also supported by the Cooperative Agreement NU58DP007114, funded by the Centers for Disease Control and Prevention. Its contents are solely the responsibility of the authors and do not necessarily represent the official views of the Centers for Disease Control and Prevention or the Department of Health and Human Services. The authors thank the other investigators, the staff, and the participants of the MESA study for their valuable contributions. A full list of participating MESA investigators and institutes can be found at http://www.mesa-nhlbi.org.

