## Supplemental Methods for "The Association between Proteomic Aging Clocks and the Risk of Cancer in Midlife Individuals"

*The ARIC study*

Study population

The ARIC study (RRID: SCR_021769) is a prospective cohort initiated in 1987 [1, 2]. In 1987-1989, 15,792 mostly White and Black volunteers aged 45-64 were recruited from four study centers - Maryland; Minnesota; Mississippi; and North Carolina. Participants in Minnesota and Maryland were primarily White, and the recruitment of Mississippi was restricted to Black residents. ARIC study was approved by institutional review boards at each participating center, and all study participants provided written informed consent. Thus far, ten study visits have been completed [1]. Additionally, ARIC participants underwent follow-up via telephone calls annually in 1987-2012 and semi-annually after 2012, with response rates of 90%-99% for the annual follow-up calls and 83%-90% for semi-annual follow-up calls among living participants who have not withdrawn consent to be contacted [2].

Ascertainment of cancer cases

Incident cancer cases were ascertained through 2015 via linkage with state cancer registries in Maryland, Minnesota, Mississippi, and North Carolina. These records were supplemented by the abstraction of medical records and hospital discharge codes [2]. In this study, obesity-related cancers include oropharynx, esophagus, postmenopausal breast, liver, gallbladder, pancreatic, stomach, kidney, colorectal, endometrial, and ovarian, and lethal prostate cancers [3]. Smoking-related cancers include head and neck, esophagus, lung, laryngeal, bladder, liver, pancreatic, stomach, kidney, colorectal and cervical cancers [4].

Blood collection

The ARIC protocol for blood sample collection, processing, and storage was designed to minimize the spontaneous biochemical reactions after blood collection and is consistent with recommended practice for proteomic data analysis in epidemiological studies [5-7]. Briefly, after venipuncture, blood samples were put immediately in an ice water bath. Centrifugation was then performed within 10 min after venipuncture at room temperature (15-25 °C). After centrifugation, the aliquots were stored at −80 °C within 90 min from venipuncture and were never thawed before this analysis.

Protein measurement and quality control

For this study, we used proteins measured in EDTA-plasma samples collected at Visit 2 (1990-92) among midlife participants (aged 46-70 years) and at Visit 5 (2011-13) among late-life participants (aged 66-90). Samples were analyzed using a Slow Off-rate Modified Aptamers (SOMAmer)--based capture array called SomaScan® 5k by Somalogic, Inc. (Boulder, CO, USA) [8-11]. The SomaScan® platform uses single-stranded DNA-based aptamers to capture conformational protein epitopes. The details of the SomaScan® assay and the data normalization process have been described previously [7, 11, 12]. Among the 5,284 available aptamer, 329 aptamers with a Bland-Altman coefficient of variation (CVBA) greater than 50% or a variance of less than 0.01 on the log scale, or binding to mouse Fc-fusion, contaminants, or non-proteins were excluded [13]. After exclusion, 4,955 aptamers (corresponding to 4,712 proteins) were included. The CVBA for split samples was 6% at Visit 2 and 7% at Visit 5. Protein concentrations were expressed in relative fluorescent units (RFU) and were log2-transformed to correct for skewness.

Assessment of other participants' characteristics

Other characteristics of interest included demographic and lifestyle characteristics and medical history, namely chronological age, sex, race, study center, education, smoking status, pack-years of smoking, alcohol intake, body mass index (BMI), hormone replacement therapy (for females only), aspirin use, diabetes status, and estimated glomerular filtration rate (eGFR). At Visit 1, participants reported education attainment [14]. At each visit, participants reported information on smoking history, alcohol intake, and use of medications and underwent a physical exam that included height and weight. Diabetes mellitus was defined as fasting glucose ≥126 mg/dL, non-fasting glucose ≥200 mg/dL, treatment for diabetes mellitus, or self-reported physician diagnosis of diabetes. The detailed procedures for assessing pack-years of smoking have been published.[15] eGFR was calculated based on serum creatinine and cystatin C and incorporated age and sex [16].

*The MESA study*

The MESA study is a prospective cohort of White, Black, Hispanic, and Chinese participants. At Exam 1 (2000-02), 6,814 participants aged 45-84 years were recruited from six field centers: Baltimore, Maryland; Chicago, Illinois; Forsyth County, North Carolina; Los Angeles, California; New York, New York; and Minneapolis, Minnesota. Each field center recruited approximately equal numbers of female and male participants from two or more of the racial/ethnic groups [17]. Details of the MESA study are available at the MESA website (https://www.mesa-nhlbi.org). MESA participants are contacted every 9-12 months to assess events including hospitalizations, which led to requests for hospitalization records. The hospitalization records contained International Classification of Diseases, 9th Revision (ICD-9) diagnosis codes relevant to that hospitalization. ICD-9 codes related to cancer (140-209) were used to determine incident cancer until 2018 [18]. MESA has measured around 7,000 proteins in the blood samples collected at Exam 1 using SomaScan® 7k, which includes all the proteins measured in ARIC.

*Statistical analysis*

PACs were constructed using R (version 4.1.2, package "glmnet") and all the other analyses were performed using SAS version 9.4 (SAS Institute Inc, Cary, NC). Statistical significance was considered if a two-sided p-value <0.05.

We constructed CaPACs among midlife participants who remained cancer-free (called here cancer-free participants) in 2015 (the end of follow-up for cancer in ARIC). In brief, among 10,834 White and Black midlife participants who were at risk for cancer at baseline (Visit 2, 1990-92), 7,487 remained cancer-free, while 3,347 developed cancer through 2015 (**Figure 1**). Among those 7,487 cancer-free participants, we randomly selected two-thirds of participants (N = 4,991) and used them as the training set. In the training set, CaPAC0 was trained against chronological age by applying elastic net regression (alpha=0.5 and lambda value was selected based on 10-fold cross-validation) [19-21]. Elastic net regression selected 1,282 aptamers with non-zero weights and CaPAC0 was constructed as a weighted sum of protein: $\beta_0 + \sum_{i=1}^{1,282} \beta_i \times aptamer_i$, where $aptamer_i$ represents the level of *ith* aptamers, $\beta_0$, the intercept, and $\beta_I$, non-zero weights [22]. In our previous study, we explored to train PACs with different alpha values. All these PACs were highly correlated with each other, so we decided to use an alpha value of 0.5, which was used in all previous studies [21].

To capture the effect of CaPAC0 independent of chronological age, we estimated age acceleration for CaPAC0 (called here CaPAA0) by regressing CaPAC0 on chronological age. We compared the distributions of characteristics among cancer-free participants (test set) and those who developed cancer during follow-up. We used Cox proportional hazards regression to estimate hazard ratios (HRs) and 95% confidence intervals (CIs) for the risk of overall cancer, obesity-related cancers, smoking-related cancers, and most common individual cancers, i.e., breast (postmenopausal breast cancer in this study), prostate, lung, and colorectal, in relation to per 5-year increase in CaPAA0. For each participant, total person-years was determined from Visit 2 date (baseline in this study) until cancer diagnosis, death, loss to follow-up, or administrative censoring on December 31, 2015, whichever occurred first. Proportional hazards assumption was examined by modeling an interaction of age acceleration and follow-up time and was not violated in any regression models. To account for the exclusion of the participants in training set who were used to construct CaPAC0, we applied a case-cohort study design employing Barlow’s method [23]. In the case-cohort analysis, we created a “sub-cohort”, which has the same distribution of cancer as the baseline study population (i.e., those 10,834 participants at risk for cancer). For example, in the analysis of overall cancer risk, the sub-cohort comprised participants who remained cancer-free (test set) and one-third randomly selected participants who developed cancer during follow-up, since the training and test sets split was 2:1. Cancer cases not included in the sub-cohort were incorporated into the analysis at the time of their cancer diagnosis. We modeled CaPAA0 as a continuous variable because no nonlinear association was observed when applying cubic splines. We adjusted for chronological age, sex, race-center (a five-category variable: Black participants from Mississippi; Black participants from any other centers; White participants from Maryland; White participants from North Carolina; and White participants from Minnesota), education, BMI, smoking status, pack-years of smoking, alcohol intake, aspirin use, hormone replacement therapy (females only) diabetes and eGFR (fully-adjusted model). In this study, we report results for a fully-adjusted model because the HRs for cancer risk were very similar in the model adjusted for demographic factors (chronological age, sex, race-center) and the fully-adjusted model. To exclude the effect of undiagnosed cancer, in a sensitivity analysis, we excluded participants who developed cancer within two years of blood collection and within four years of blood collection and then replicated our analysis for overall, smoking-related, lung and colorectal cancers (these are the cancers we found significant associations with age acceleration in our study).

In MESA, after excluding participants with self-reported history of cancer at Exam 1 (2000-02), we computed CaPAC0 in 3,893 MESA participants who were 46-70 years old (the same age range as participants in ARIC). Among those 3,893 participants, 456 developed cancer through 2018. We examined the association between age acceleration and cancer risk until 2018. In an additional analysis, we applied CaPAC0 to all MESA participants (aged 45-84 years ) at Exam 1 without a history of cancer (N=5,330) and examined its association with incident cancer (N=678).

In ARIC and MESA, we conducted subgroups analyses by stratifying the associations with the risk of overall cancer, obesity-related cancers, and smoking-related cancers by gender, race, and smoking status; it is known that males [24] and smokers [25] might age faster and smokers and Black individuals had a higher incidence of several cancer types [26]. In ARIC, we conducted three additional analyses: 1) we examined the association with smoking-related cancers among current and former smokers stratified by alcohol intake as individual who smoke are likely to be alcohol drinkers [27]; 2) We explored whether the associations of CaPAA0 with smoking-related cancers risk were driven by the most common cancers;

and 3) we examined association between age acceleration and lung cancer risk stratified by smoking status because smoking is a strong risk factor for lung cancer. We did not conduct these analyses in MESA because of the limited number of cancer cases.

We also created two additional CaPACs and examined their association with cancer risk. First, we aimed to develop a CaPAC including ~20 aptamers (CaPAC1) to test if its associations with cancer risk are similar to those for the original CaPAC comprising 1282 aptamers. We constructed CaPAC1, comprising 21 aptamers, by training it against chronological age. Second, we selected the top 40% of proteins with the largest change from midlife to late life. We constructed CaPAC2 by training those selected proteins against chronological age at Visit 2. Elastic net regression finally selected 420 aptamers for CaPAC2. Moreover, we computed Lehallier's PAC and examined its association with cancer risk. We estimated weights for Lehallier's PAC instead of using the published weights, because ARIC did not measure all the 491 aptamers reported in Lehallier [2020] [28]. We re-estimated weights for Lehallier's PAC by applying Ridge regression in the training set. In our previous study, we found that a published PAC calculated using published weights and ARIC weights showed similar performance [21]. Finally, we computed BioAge developed in Levine [2013] [29]. BioAge was originally developed using 10 clinical markers that represent the decline in age-related physiological functioning and susceptibility to disease in old age [29]. These 10 markers include systolic blood pressure (SBP), total cholesterol, fasting glucose, cytomegalovirus infection (CMV), C-Reactive Protein (CRP), serum creatinine, blood urea nitrogen (BUN), alkaline phosphatase, albumin, and peak flow measurement. In ARIC, we constructed BioAge based on SBP, total cholesterol, fasting glucose, CRP, serum creatinine, alkaline phosphatase, and peak flow measurement measured at Visit 2 (1990-92), as well as BUN and albumin measured at Visit 1 (1987-89). CMV was not measured in ARIC. BioAge in ARIC was computed using R package "BioAge" [30].
