## Supplemental Tables for "The Association between Proteomic Aging Clocks and the Risk of Cancer in Midlife Individuals"

Supplemental Table 1. Functions of the top 20 proteins with the largest absolute weights in CaPAC0[a]

| | Seqid | Weights | UniProt ID | Protein | Gene | Functions[b] |
|---|---|---|---|---|---|---|
| 1 | SeqId_16890_37 | 2.22886787 | Q8N6G6 | ADAMTS-like protein 1 | ADAMTSL1 | May have important functions in the extracellular matrix |
| 2 | SeqId_14136_234 | -1.980828084 | Q9NPY3 | Complement component C1q receptor | CD93 | Plays a role in various physiological processes including inflammation, phagocytosis, and cell adhesion |
| 3 | SeqId_3362_61 | 1.900352521 | Q9BU40 | Chordin-like protein 1 | CHRDL1 | May play a role in topographic retinotectal projection and in the regulation of retinal angiogenesis in response to hypoxia |
| 4 | SeqId_15640_54 | 1.78847103 | Q01995 | Transgelin | TAGLN | Acts as a tumor suppressor, and the loss of its expression is an early event in cell transformation and the development of some tumors, coinciding with cellular plasticity. |
| 5 | SeqId_8956_96 | 1.765511275 | Q96GP6 | Scavenger receptor class F member 2 | SCARF2 | Mediates homophilic and heterophilic interactions |
| 6 | SeqId_3045_72 | 1.671734852 | P21246 | Pleiotrophin | PTN | Regulates processes like cell proliferation, cell survival, cell growth, cell differentiation and cell migration |
| 7 | SeqId_6392_7 | 1.627165568 | O76076 | WNT1-inducible-signaling pathway protein 2 | CCN5 | This protein is encoded by a gene that may be downstream in the WNT1 signaling pathway that is relevant to malignant transformation. The expression of this gene in colon tumors is reduced while the other two WISP members are overexpressed in colon tumors. |
| 8 | SeqId_9793_145 | -1.383408273 | Q8TDY8 | Immunoglobulin superfamily DCC subclass member 4 | IGDCC4 | Belongs to the immunoglobulin superfamily. |
| 9 | SeqId_8974_172 | -1.375527134 | P39059 | Collagen alpha-1(XV) chain | COL15A1 | Stabilizes microvessels and muscle cells, both in heart and in skeletal muscle |
| 10 | SeqId_10514_5 | 1.362832231 | P41222 | Prostaglandin-H2 D-isomerase | PTGDS | Functions as a neuromodulator as well as a trophic factor in the central nervous system |
| 11 | SeqId_3009_3 | -1.26655331 | Q03167 | Transforming growth factor beta receptor type 3 | TGFBR3 | Decreased expression of this receptor has been observed in various cancers. |
| 12 | SeqId_3168_8 | 1.188397617 | Q9UNA0 | A disintegrin and metalloproteinase with thrombospondin motifs 5 | ADAMTS5 | Plays an important role in connective tissue organization, development, inflammation, arthritis, and cell migration |
| 13 | SeqId_4374_45 | 1.14295554 | Q99988 | Growth/differentiation factor 15 | GDF15 | Acts as a pleiotropic cytokine and is involved in the stress response program of cells after cellular injury; Increased protein levels are associated with disease states such as |

| | | | | | | |
|---|---|---|---|---|---|---|
| | | | | | | tissue hypoxia, inflammation, acute injury and oxidative stress |
| 14 | SeqId_12417_46 | 1.094384467 | Q9Y3C4 | EKC/KEOPS complex subunit TPRKB | TPRKB | Component of the EKC/KEOPS complex that is required for the formation of a threonylcarbamoyl group on adenosine at position 37 ($t_6$A37) in tRNAs that read codons beginning with adenine |
| 15 | SeqId_9525_1 | -1.057668956 | Q13308 | Inactive tyrosine-protein kinase 7 | PTK7 | involves in Wnt signaling pathway and plays a role in multiple cellular processes including polarity and adhesion |
| 16 | SeqId_3607_71 | 1.040027712 | Q9UBP4 | Dickkopf-related protein 3 | DKK3 | This protein is encoded by a gene that may function as a tumor suppressor |
| 17 | SeqId_5740_17 | -1.025055944 | Q9Y6N7 | Roundabout homolog 1 | ROBO1 | Receptor for SLIT1 and SLIT2 that mediates cellular responses to molecular guidance cues in cellular migration, including axonal navigation at the ventral midline of the neural tube and projection of axons to different regions during neuronal development |
| 18 | SeqId_12630_8 | 1.020866882 | P53365 | Arfaptin-2 | ARFIP2 | Plays a role in constitutive metalloproteinase (MMP) secretion from the trans Golgi network |
| 19 | SeqId_13114_50 | 0.97456564 | P51884 | Lumican | LUM | May regulate collagen fibril organization and circumferential growth, corneal transparency, and epithelial cell migration and tissue repair |
| 20 | SeqId_2677_1 | -0.969095072 | P00533 | Epidermal growth factor receptor | EGFR | a cell surface protein that binds to epidermal growth factor, inducing receptor dimerization and tyrosine autophosphorylation leading to cell proliferation |

[a] CaPAC was constructed among participants who remained cancer-free (cancer-free participants, training set) in ARIC.

[b] Fuctions of proteins documented in GeneCards (https://www.genecards.org/).

Supplemental Table 2. Association between age acceleration for CaPAC0 (CaPAA0)[a] and risk of smoking-related cancers after excluding common cancers (lung and colorectal), ARIC (1992-2015)

| | HR (95% CI)[b] per 5-years of CaPAA0 |
|---|---|
| Smoking-related cancers[c] | 1.30 (1.17, 1.45) |
| Smoking-related cancers after excluding those with lung cancer | 1.20 (1.05, 1.38) |
| Smoking-related cancers after excluding those with colorectal cancer | 1.30 (1.15, 1.47) |

[a] CaPAC0 was constructed among participants who remained cancer-free (cancer-free participants, training set) in ARIC. CaPAA0 was calculated as residuals after regressing CaPAC0 on chronological age.

[b] The model was adjusted for chronological age, sex, race-center, education, body mass index (BMI), smoking status, pack-years of smoking, alcohol intake, aspirin use, hormone replacement therapy (among females), diabetes and eGFR. HRs were calculated using a case-cohort study design to account for the exclusion of training set used to develop CaPAC0.

[c] Smoking-related cancers include head and neck, esophagus, lung, laryngeal, bladder, liver, pancreatic, stomach, kidney, colorectal and cervical cancers.

Supplemental Table 3. Association of age acceleration for CaPAC0 (CaPAA0)[a] with the risk of overall, smoking-related, lung, and colorectal cancers among participants after excluding those developed cancer within two and four years of blood collection; ARIC (1990-2015)

| | HR (95% CI)[b] per 5 years of CaPAA0 | | |
|---|---|---|---|
| | Among all participants (after excluding training set) | Among participants (after excluding training set) after excluding those developed cancer within two years of blood collection | Among participants (after excluding training set) after excluding those developed cancer within four years of blood collection |
| Overall cancer | 1.08 (1.00, 1.18) | 1.07 (1.00, 1.15) | 1.08 (1.00, 1.16) |
| Smoking-related cancers[c] | 1.30 (1.17, 1.45) | 1.31 (1.17, 1.46) | 1.34 (1.19, 1.50) |
| Lung cancer | 1.54 (1.28, 1.86) | 1.47 (1.22, 1.77) | 1.51 (1.24, 1.84) |
| Colorectal cancer | 1.31 (1.05, 1.63) | 1.31 (1.04, 1.66) | 1.31 (1.05, 1.69) |

[a] CaPAC0 was constructed among participants who remained cancer-free (cancer-free participants, training set) in ARIC. CaPAA0 was calculated as residuals after regressing CaPAC0 on chronological age.

[b] The model was adjusted for chronological age, sex, race-center, education, body mass index (BMI), smoking status, pack-years of smoking, alcohol intake, aspirin use, hormone replacement therapy (among females), diabetes and eGFR. HRs were calculated using a case-cohort study design to account for the exclusion of training set used to develop CaPAC0.

[c] Smoking-related cancers include head and neck, esophagus, lung, laryngeal, bladder, liver, pancreatic, stomach, kidney, colorectal and cervical cancers.

Supplemental Table 4. Association between age acceleration for CaPAC0 (CaPAA0)[a] and cancer risk in participants with different ages, MESA (2000-2018)

| | Participants aged 46-70 years (the same age range as in ARIC) | | Participants aged 45-84 years | |
|---|---|---|---|---|
| | N of cases[b] | HR (95% CI)[c] per 5-year in CaPAA0 | N of cases[b] | HR (95% CI)[c] per 5-year in CaPAA0 |
| Overall cancer | 456 | 1.23 (1.03, 1.47) | 678 | 1.22 (1.06, 1.40) |
| Obesity-related cancers[d] | 149 | 1.20 (0.88, 1.63) | 222 | 1.20 (0.94, 1.52) |
| Smoking-related cancers[d] | 146 | 1.54 (1.13, 2.09) | 232 | 1.45 (1.15, 1.83) |
| Prostate cancer | 92 | 1.28 (0.86, 1.90) | 138 | 1.44 (1.06, 1.96) |
| Lung cancer | 55 | 1.94 (1.18, 3.17) | 84 | 1.71 (1.25, 2.33) |
| Postmenopausal breast cancer | 34 | 0.87 (0.46, 1.68) | 47 | 0.87 (0.52, 1.48) |
| Colorectal cancer | 33 | 0.78 (0.38, 1.53) | 60 | 0.92 (0.57, 1.48) |

[a] CaPAC0 was constructed among participants who remained cancer-free (cancer-free participants, training set) in ARIC. CaPAA0 was calculated as residuals after regressing CaPAC0 on chronological age.

[b] Participants with missing covariates of interest were excluded from the analysis.

[c] The model was adjusted for chronological age, sex, race-center, education, body mass index (BMI), smoking status, pack-years of smoking, alcohol intake, aspirin use, hormone replacement therapy (among females), diabetes and eGFR.

[d] Obesity-related cancers include oropharynx, esophagus, postmenopausal breast, liver, gallbladder, pancreatic, stomach, kidney, colorectal, endometrial, and ovarian, and lethal prostate cancers. Smoking-related cancers include head and neck, esophagus, lung, laryngeal, bladder, liver, pancreatic, stomach, kidney, colorectal and cervical cancers.

Supplemental Table 5. Association between age acceleration for CaPACs[a] and cancer risk among participants aged 46-70 in ARIC (1990-2015) and in MESA (2000-2018)

| | ARIC (after excluding the training set, aged 46-70 years) | | | MESA (aged 46-70 years) | | |
|---|---|---|---|---|---|---|
| | HR (95% CI)[b] per 5-years of age acceleration | | | HR (95% CI)[c] per 5-years of age acceleration | | |
| | CaPAC0 | CaPAC1 | CaPAC2 | CaPAC0 | CaPAC1 | CaPAC2 |
| Overall cancer risk | 1.08 (1.00, 1.18) | 1.22 (1.03, 1.43) | 0.98 (0.91, 1.05) | 1.23 (1.03, 1.47) | 1.94 (1.39, 2.72) | 1.21 (1.02, 1.45) |
| Obesity-related cancers[c] risk | 1.04 (0.93, 1.16) | 1.19 (0.93. 1.53) | 0.94 (0.84, 1.06) | 1.20 (0.88, 1.63) | 2.94 (1.67, 5.16) | 1.37 (1.01, 1.86) |
| Smoking-related cancers[c] risk | 1.30 (1.17, 1.45) | 1.77 (1.39, 2.25) | 1.22 (1.08, 1.37) | 1.54 (1.13, 2.09) | 3.46 (1.98, 6.03) | 1.56 (1.14, 2.13) |
| Prostate cancer risk | 0.92 (0.78, 1.10) | 0.67 (0.46, 1.00) | 1.05 (0.89, 1.26) | 1.28 (0.86, 1.90) | 1.05 (0.46, 2.36) | 1.13 (0.75, 1.69) |
| Lung cancer risk | 1.54 (1.28, 1.86) | 2.33 (1.58, 3.44) | 1.50 (1.24, 1.84) | 1.94 (1.18, 3.17) | 3.18 (1.23, 8.20) | 1.52 (0.92, 2.53) |
| Postmenopausal-breast cancer risk | 0.90 (0.75, 1.09) | 0.97 (0.64, 1.46) | 0.72 (0.60, 0.90) | 0.87 (0.46, 1.68) | 1.88 (0.52, 6.76) | 1.11 (0.58, 2.11) |
| Colorectal cancer risk | 1.31 (1.05, 1.63) | 1.93 (1.16, 3.21) | 1.17 (0.93, 1.50) | 0.78 (0.38, 1.53) | 1.89 (0.55, 6.54) | 1.21 (0.63, 2.33) |

[a] CaPAC0 was constructed among participants who remained cancer-free (cancer-free participants, training set) in ARIC. CaPAC1 was constructed similarly as CaPAC1 but limited to a small group of proteins (N=21). CaPAC2 consisted of 420 proteins which were selected by elastic net regression from the top 40% of proteins with the largest change from midlife to late life in ARIC. Age acceleration was calculated as residuals after regressing CaPACs on chronological age.

[b] The model was adjusted for chronological age, sex, race-center, education, body mass index (BMI), smoking status, pack-years of smoking, alcohol intake, aspirin use, hormone replacement therapy (among females), diabetes and eGFR. HRs in ARIC were calculated using a case-cohort study design.

[c] Obesity-related cancers include oropharynx, esophagus, postmenopausal breast, liver, gallbladder, pancreatic, stomach, kidney, colorectal, endometrial, and ovarian, and lethal prostate cancers. Smoking-related cancers include head and neck, esophagus, lung, laryngeal, bladder, liver, pancreatic, stomach, kidney, colorectal and cervical cancers.

Supplemental Table 6. Association between age acceleration for CaPAC0[a] and Lehallier's PAC and cancer risk among participants aged 46-70 in ARIC (1990-2015) and in MESA (2000-2018)

| | ARIC (after excluding the training set, aged 46-70 years) | | MESA (aged 46-70 years) | |
|---|---|---|---|---|
| | HR (95% CI)[b] per 5-years of age acceleration | | HR (95% CI) per 5-years of age acceleration | |
| | CaPAC0[a] | Lehallier's PAC | CaPAC0 | Lehallier's PAC |
| Overall cancer risk | 1.08 (1.00, 1.18) | 1.06 (0.96, 1.15) | 1.23 (1.03, 1.47) | 1.20 (1.01, 1.41) |
| Obesity-related cancers[c] risk | 1.04 (0.93, 1.16) | 0.98 (0.89, 1.09) | 1.20 (0.88, 1.63) | 1.03 (0.78, 1.38) |
| Smoking-related cancers[c] risk | 1.30 (1.17, 1.45) | 1.24 (1.11, 1.37) | 1.54 (1.13, 2.09) | 1.20 (0.90, 1.61) |
| Prostate cancer risk | 0.92 (0.78, 1.10) | 0.91 (0.77, 1.07) | 1.28 (0.86, 1.90) | 1.46 (1.00, 2.12) |
| Lung cancer risk | 1.54 (1.28, 1.86) | 1.54 (1.28, 1.84) | 1.94 (1.18, 3.17) | 1.40 (0.86, 2.28) |
| Postmenopausal-breast cancer risk | 0.90 (0.75, 1.09) | 0.94 (0.79, 1.12) | 0.87 (0.46, 1.68) | 1.07 (0.58, 1.95) |
| Colorectal cancer risk | 1.31 (1.05, 1.63) | 1.14 (0.92, 1.42) | 0.78 (0.38, 1.53) | 0.68 (0.36, 1.27) |

[a] CaPAC0 was constructed among participants who remained cancer-free (cancer-free participants, training set) in ARIC. Age acceleration was calculated as residuals after regressing PAC on chronological age.

[b] The model was adjusted for chronological age, sex, race-center, education, body mass index (BMI), smoking status, pack-years of smoking, alcohol intake, aspirin use, hormone replacement therapy (among females), diabetes and eGFR. HRs in ARIC were calculated using a case-cohort study design.

[c] Obesity-related cancers include oropharynx, esophagus, postmenopausal breast, liver, gallbladder, pancreatic, stomach, kidney, colorectal, endometrial, and ovarian, and lethal prostate cancers. Smoking-related cancers include head and neck, esophagus, lung, laryngeal, bladder, liver, pancreatic, stomach, kidney, colorectal and cervical cancers.

Supplemental Table 7. Association of age acceleration for Klemera-Doubal biological age (KDM-BA)[a] and age acceleration for CaPAC0[a] (CaPAA0) with cancer risk when including age acceleration for KDM-BA and CaPAA0 in the same model, ARIC (1990-2015)

| | HR (95% CI)[b] per 5-years of age acceleration |
|---|---|
| Age acceleration for KDM-BA | 1.32 (1.08, 1.60) |
| Age acceleration for CaPAC0 (CaPAA0) | 1.44 (1.20, 1.74) |

[a] KDM-BA was constructed using clinical and physiological markers in Levine [2013]. CaPAC0 was constructed among participants who remained cancer-free (cancer-free participants, training set) in ARIC.

[b] The model was adjusted for chronological age, sex, race-center, education, BMI, smoking status, pack-years of smoking, alcohol intake, aspirin use, diabetes and eGFR.
